# Persistent Racial Inequities in Acute Kidney Injury Among U.S. Hospitalizations: A Nationwide Cohort Analysis

**DOI:** 10.64898/2026.03.24.26349246

**Authors:** Brent Tai, Chijioke Okonkwo

## Abstract

**Background:** Acute kidney injury (AKI) is a major contributor to morbidity, mortality, and healthcare utilization among hospitalized adults. Long-standing racial and ethnic inequities in U.S. healthcare—including unequal access to care, neighborhood disadvantage, and other structural factors—are known to influence kidney health, yet national data describing how these inequities manifest in AKI remain limited.

**Methods:** We conducted a retrospective, cross-sectional analysis of the 2022 National Inpatient Sample. AKI was identified using ICD-10-CM codes N17.x, and race/ethnicity followed HCUP categories. Descriptive analyses compared characteristics across groups. Survey-weighted logistic regression estimated adjusted odds of developing AKI, in-hospital mortality among AKI patients, and dialysis use, adjusting for demographics, payer, and comorbidities. Age-specific predicted AKI probabilities were derived from the adjusted model.

**Results:** AKI prevalence ranged from 15% to 23% across racial and ethnic groups. After adjustment, Black (OR 1.34), Native American (OR 1.08), and Other patients (OR 1.07) had higher odds of AKI, whereas Asian/Pacific Islander (OR 0.94) and Hispanic (OR 0.98) had slightly lower or similar odds. Among AKI hospitalizations, mortality was modestly lower for Black and Hispanic patients relative to White patients and higher for Asian/Pacific Islander and Native American patients. All non-White groups had higher odds of dialysis use. Age-specific curves showed persistent risk differences across adulthood.

**Conclusions:** Substantial racial disparities in AKI incidence, mortality, and dialysis use persisted after adjustment, reflecting broader structural inequities. Addressing these gaps will require both targeted clinical strategies and policy interventions focused on upstream determinants.

## Introduction

Acute kidney injury (AKI) is a common and clinically significant complication among hospitalized adults, associated with higher mortality, prolonged length of stay, increased healthcare costs, and long-term progression to chronic kidney disease (CKD) [3,12,23]. Despite advances in recognition and supportive management, AKI remains a major contributor to inpatient morbidity and resource utilization across the United States. Understanding contemporary epidemiologic patterns is essential given the long-standing and persistent influence of structural racism—through differential exposure to risk factors, unequal access to preventive and acute care, and variation in hospital quality—that continues to shape patterns of kidney disease in the United States.

Racial and ethnic disparities in kidney disease are well documented, with minority populations experiencing disproportionate rates of CKD, end-stage kidney disease, and AKI [14–15]. These inequities are shaped by complex and interrelated factors—including structural racism, differential access to primary and specialty care, environmental exposures, and socioeconomic disadvantage [17–18]. However, existing studies on racial disparities in AKI often rely on older data, focus on limited outcomes, or do not fully account for socioeconomic gradients and hospital-level factors. Furthermore, little is known about whether racial differences in AKI risk and severity vary across the adult lifespan or remain consistent at all ages.

Socioeconomic status is another important determinant of health, yet prior work suggests that income alone does not fully explain the racial inequalities observed in kidney disease [19,21–22]. Minority patients may face compounding structural barriers in addition to financial disadvantage, including residential segregation [1,13], reduced healthcare access [4], and variability in hospital quality [2]. Understanding how income and race intersect to shape AKI outcomes is essential for developing targeted, equitable prevention strategies.

In addition to clinical outcomes, disparities in the use of kidney replacement therapy and post-acute care services have been reported across racial and ethnic groups [24]. These patterns have broad implications for healthcare costs, resource allocation, and long-term recovery. Nevertheless, contemporary data describing how these disparities manifest in AKI-related hospitalization at a national level remain limited.

To address these gaps, we aimed to generate a contemporary national profile of racial disparities in acute kidney injury (AKI) using the 2022 National Inpatient Sample. Our objectives were threefold: (1) to compare AKI prevalence and unadjusted clinical outcomes across racial and ethnic groups; (2) to quantify adjusted associations between race and AKI development, in-hospital mortality, and dialysis use; and (3) to model age-specific predicted AKI risk to determine whether disparities vary across the adult lifespan. This integrated approach allows us to characterize inequities in both absolute burden and adjusted risk, providing one of the most comprehensive assessments of racial disparities in AKI.

## Materials and Methods

### Study Design and Data Source

We conducted a retrospective, cross-sectional study using the 2022 National Inpatient Sample (NIS), the largest all-payer inpatient database in the United States [7]. The NIS is produced by the Healthcare Cost and Utilization Project (HCUP) and captures approximately 35 million annual hospitalizations, which can be weighted to generate nationally representative estimates. All analyses incorporated survey weights, clusters, and strata to account for the complex sampling design. Consistent with HCUP guidance, we did not exclude transfer hospitalizations, elective admissions, or hospice discharges, as removing these records may bias national estimates. All discharge dispositions provided in the NIS were retained for analysis. Elective versus non-elective admission type was not used as an exclusion criterion. This study used publicly available, de-identified data and was exempt from institutional review board review by BayCare Health Care.

### Study Population

All hospitalized adults aged ≥18 years were eligible. Acute kidney injury (AKI) was defined using ICD-10-CM codes N17.x across all diagnosis fields. Race/ethnicity followed HCUP categories: Asian/Pacific Islander (API), Black, Hispanic, Native American, Other, and White. HCUP defines “Other” as non-White, non-Black, non-Hispanic, non-API, non-Native American. We excluded hospitalizations with missing race data and pregnancy-related admissions.

### Variables and Definitions

Baseline variables included age, sex, primary payer, ZIP code–level median household income quartile, and comorbidities: CKD, heart failure, diabetes, hypertension, obesity, COPD, liver disease, and sepsis. Race/ethnicity was missing for 3.2% of hospitalizations; these records were excluded per prespecified criteria because race was a primary exposure variable. Other variables had low missingness (<2%), and missing values were handled using complete-case analysis, consistent with HCUP recommendations. No imputation was performed. The final analytic cohort therefore included only observations with complete race data and complete covariate information required for regression modeling.

Race/ethnicity followed HCUP classifications, which reflect self-reported or administratively recorded categories using standardized coding conventions. Median household income was defined by ZIP code–level income quartile, as provided in HCUP. Income quartile was included in descriptive tables to characterize socioeconomic distribution across racial and ethnic groups and AKI status. In regression models, income quartile was included as an adjustment variable to account for socioeconomic differences that may influence AKI risk and outcomes. No mediation analyses were performed. Income was also examined as a potential effect modifier descriptively, but no interaction terms were included in final models.

### Clinical Outcomes

Primary outcomes were in-hospital mortality and dialysis use among AKI patients. Secondary outcomes included mechanical ventilation, shock, cardiac arrest, length of stay (LOS), total hospital charges, and discharge disposition (home, home health care, SNF, transfer, AMA). LOS and charges were survey-weighted; charges were log-transformed for modeling and visualization.

### Statistical Analysis

Descriptive statistics used weighted means and percentages, with standardized mean differences to compare groups. Three survey-weighted logistic regression models evaluated associations between race and a) development of AKI, b) in-hospital mortality among AKI patients, c) dialysis use among AKI patients. All models adjusted for age, sex, payer, CKD, heart failure, diabetes, hypertension, obesity, COPD, liver disease, and sepsis (White reference group). Age-specific predicted probabilities of AKI were estimated from the adjusted model, with covariates fixed at reference values, and plotted as risk differences versus White patients. Analyses were performed in R 4.3 using the survey, srvyr, tidyverse, broom, and ggeffects packages.

## Results

### Baseline Characteristics

All percentages, means, and rates reported in Tables and Figures represent survey-weighted national estimates unless otherwise specified. Baseline demographic and clinical characteristics stratified by race are presented in Table 1. White patients were older on average (mean age 62.1 years) and had the highest proportion aged ≥75 years (31.6%), whereas Black and Hispanic patients were younger (mean ages 53.6 and 49.9 years, respectively) and more frequently in the 18–64-year age range. CKD prevalence was highest among Black patients (25.1%) compared with White patients (18.7%). AKI prevalence varied across racial and ethnic groups, ranging from 15.3% among Hispanic patients to 22.8% among Black patients.

**Table 1.** Weighted Baseline Demographic and Clinical Characteristics of Adult Hospitalizations by Race.

| Race | Unweighted (n) | Age (mean) | Female (%) | AKI (%) | Age 18-44 (%) | Age 45-64 (%) | Age 65-74 (%) | Age $\geq 75$ (%) | CKD (%) | HF (%) | Diabetes (%) | HTN (%) | Obesity (%) | COPD (%) | Liver disease (%) | Sepsis (%) |
| --- | --- | --- | --- | --- | --- | --- | --- | --- | --- | --- | --- | --- | --- | --- | --- | --- |
| Asian/Pacific Islander | 163,368 | 55.9 | 62.6 | 16.7 | 37.9 | 20.8 | 16.5 | 24.7 | 18.8 | 15.0 | 8.5 | 6.4 | 8.1 | 4.9 | 1.1 | 11.4 |
| Black | 840,777 | 53.6 | 57.4 | 22.8 | 34.9 | 32.1 | 17.8 | 15.2 | 25.1 | 22.8 | 10.6 | 8.4 | 21.8 | 11.1 | 1.0 | 10.0 |
| Hispanic | 691,121 | 49.9 | 60.7 | 15.3 | 45.6 | 26.1 | 13.2 | 15.1 | 15.2 | 13.1 | 9.2 | 5.8 | 18.4 | 5.2 | 2.0 | 10.5 |
| Native American | 38,037 | 51.8 | 57.4 | 18.2 | 38.9 | 31.9 | 16.0 | 13.2 | 18.1 | 17.1 | 11.4 | 6.9 | 19.7 | 10.5 | 3.7 | 12.9 |

| Race | Unweighted (n) | Age (mean) | Female (%) | AKI (%) | Age 18-44 (%) | Age 45-64 (%) | Age 65-74 (%) | Age ≥75 (%) | CKD (%) | HF (%) | Diabetes (%) | HTN (%) | Obesity (%) | COPD (%) | Liver disease (%) | Sepsis (%) |
| --- | --- | --- | --- | --- | --- | --- | --- | --- | --- | --- | --- | --- | --- | --- | --- | --- |
| Other/Multiracial | 163,718 | 53.3 | 57.5 | 16.6 | 39.7 | 25.1 | 15.8 | 19.3 | 14.5 | 14.8 | 7.5 | 5.9 | 15.4 | 7.4 | 1.5 | 10.1 |
| White | 3,537,871 | 62.1 | 55.1 | 19.4 | 21.8 | 24.8 | 21.8 | 31.6 | 18.7 | 21.2 | 8.2 | 7.5 | 18.9 | 15.4 | 1.5 | 11.0 |

Table 1 summarizes demographic and clinical differences between race groups, providing context for subsequent analyses of racial disparities in AKI prevalence and associated outcomes. Unweighted counts reflect the actual number of hospitalizations in the analytic cohort. Continuous variables are presented as weighted means, and categorical variables as weighted percentages. Age categories represent the weighted proportion of patients within each group. Comorbidities—including CKD, heart failure, diabetes, hypertension, obesity, COPD, liver disease, and sepsis—were identified from ICD-10-CM diagnosis codes across all listed diagnosis fields. Acute kidney injury (AKI) was defined using ICD-10-CM codes N17.x.

Baseline characteristics by neighborhood income quartile are summarized in Table 2. Patients in the lowest income quartile were younger on average (mean age approximately 57 years) and had slightly higher prevalence of CKD and heart failure compared with those in the highest income quartile. These baseline differences in age and comorbidity profiles contextualize the unadjusted outcome patterns shown in Supplementary Table 1 and, separately, the income-stratified outcomes in Supplementary Table 2.

**Table 2.** Weighted Baseline Characteristics of Adult Hospitalizations by Income Quartile.

| Income | Unweighted (n) | Age (mean) | Female (%) | AKI (%) | Age 18-44 (%) | Age 45-64 (%) | Age 65-74 (%) | Age ≥75 (%) | CKD (%) | HF (%) | Diabetes (%) | HTN (%) | Obesity (%) | COPD (%) | Liver disease (%) | Sepsis (%) |
| --- | --- | --- | --- | --- | --- | --- | --- | --- | --- | --- | --- | --- | --- | --- | --- | --- |
| Q1 (Lowest income) | 1,635,179 | 57.1 | 56.2 | 20.2 | 29.2 | 29.2 | 19.6 | 21.9 | 19.9 | 21.3 | 9.8 | 7.7 | 20.2 | 15.5 | 1.5 | 11.1 |

| Income | Unweighted<br>(n) | Age<br>(mean) | Female<br>(%) | AKI<br>(%) | Age<br>18-44<br>(%) | Age<br>45-64<br>(%) | Age<br>65-74<br>(%) | Age<br>≥75<br>(%) | CKD<br>(%) | HF<br>(%) | Diabetes<br>(%) | HTN<br>(%) | Obesity<br>(%) | COPD<br>(%) | Liver<br>disease<br>(%) | Sepsis<br>(%) |
| --- | --- | --- | --- | --- | --- | --- | --- | --- | --- | --- | --- | --- | --- | --- | --- | --- |
| Q2 | 1,421,142 | 58.8 | 56.5 | 19.3 | 27.6 | 26.1 | 20.1 | 26.1 | 19.2 | 20.5 | 9.0 | 7.4 | 19.7 | 14.0 | 1.5 | 10.8 |
| Q3 | 1,319,809 | 59.1 | 57.1 | 18.8 | 28.2 | 24.4 | 19.7 | 27.6 | 18.9 | 19.5 | 8.4 | 7.2 | 18.9 | 11.5 | 1.5 | 10.6 |
| Q4<br>(Highest<br>income) | 1,113,439 | 60.4 | 57.4 | 17.9 | 27.7 | 22.1 | 18.9 | 31.3 | 17.9 | 17.8 | 7.0 | 6.7 | 15.8 | 8.9 | 1.4 | 10.3 |

Table 2 highlights demographic and clinical gradients across income quartiles, establishing the socioeconomic context for evaluating disparities in AKI and related clinical outcomes. Neighborhood income quartiles are based on ZIP code–level median household income classifications provided by HCUP. Unweighted counts represent the number of discharges in each quartile, while all reported summary statistics reflect survey-weighted national estimates. Age is reported as a weighted mean; categorical demographic and comorbidity measures are reported as weighted percentages. Comorbidities were derived from ICD-10-CM diagnosis codes, and AKI was identified using N17.x.

### Unadjusted Clinical Outcomes

Unadjusted clinical outcomes and discharge disposition by race are summarized in Supplementary Table 1. Dialysis use ranged from 2.1% among White patients to 6.7% among Black patients, while in-hospital mortality ranged from 2.0% among Hispanic patients to approximately 3.0% among Asian/Pacific Islander and White patients. Mechanical ventilation ranged from 3.1% among Hispanic patients to 4.8% among Native American patients, and shock occurred in approximately 1.6% to 2.4% of hospitalizations. Length of stay ranged from 4.44 days among Asian/Pacific Islander patients to 5.61 days among Black patients, and mean total hospital charges ranged from $66,331 among Native American patients to $91,549 among Other race patients. Discharge to a skilled nursing facility was most common among White patients, while leaving against medical advice occurred more often among Black and Hispanic patients. These unadjusted patterns across AKI prevalence, critical illness markers, and resource utilization measures are displayed visually in Supplementary Figure 1, which presents standardized (z-score) heatmaps of all unadjusted outcomes.

### Income-stratified outcomes

Because socioeconomic status may influence clinical outcomes, Figure 1 summarizes mortality and dialysis rates across income quartiles within each racial group. Corresponding unadjusted income-stratified outcomes are provided in Supplementary Table 2. Within-group income gradients were observed across several measures. For example, dialysis use ranged from 4.0% in the lowest income quartile to 2.5% in the highest quartile, while mean length of stay decreased from 5.19 to 4.63 days. In-hospital mortality was relatively stable across quartiles, ranging from 2.9% in the lowest quartile to 2.6% in the highest quartile. In contrast, mean hospital charges increased across quartiles, rising from $71,022 in the lowest-income neighborhoods to $82,704 in the highest-income quartile. Since the racial and ethnic groups differ markedly in their underlying sample sizes, with White and Black patients representing the majority of hospitalizations, variability in estimates for smaller groups (e.g., Native American, Asian/Pacific Islander) may appear wider due to smaller sample sizes and should therefore be interpreted cautiously.

**Figure 1.**
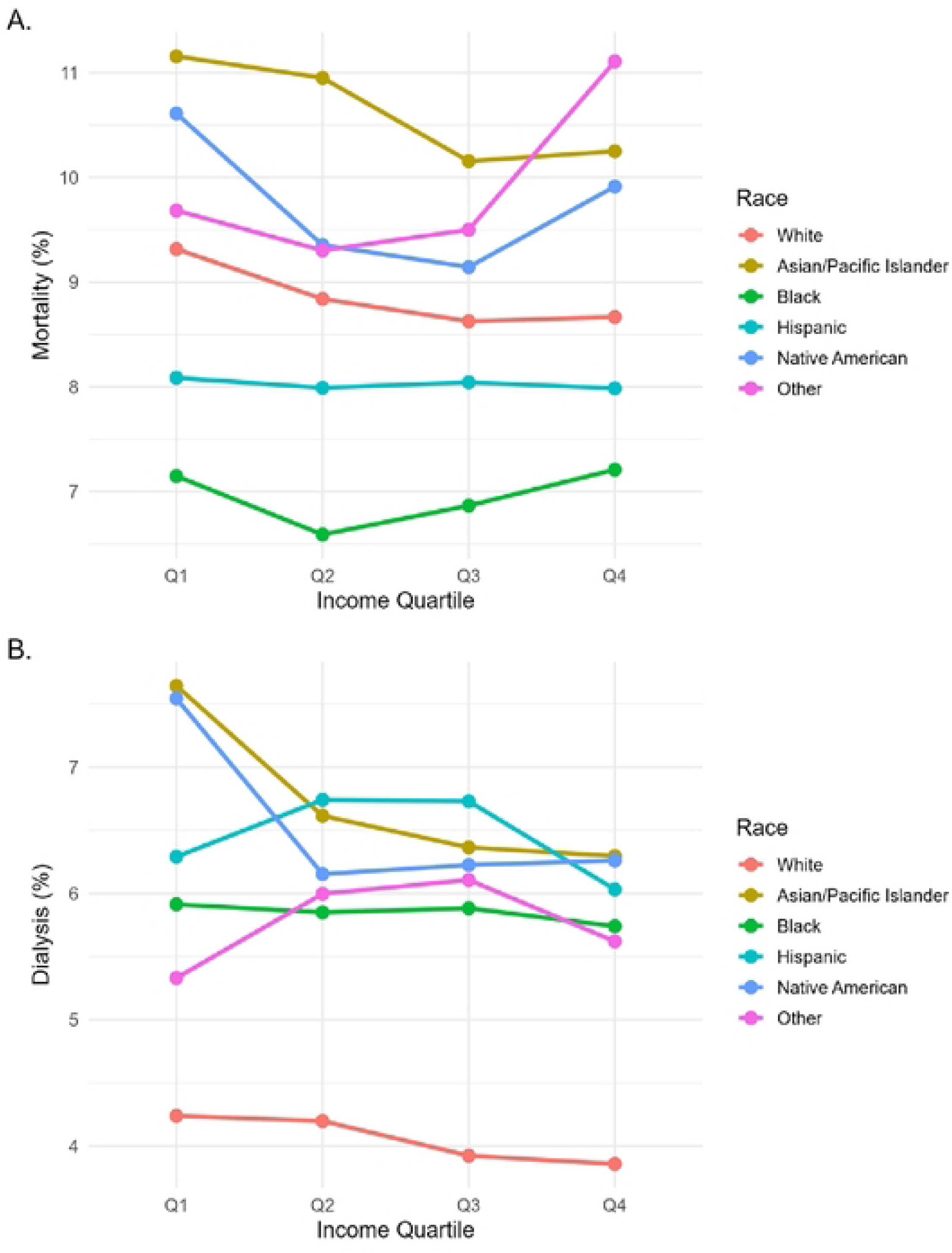
Mortality and Dialysis Rates Across Income Quartiles Within Each Racial Group.

Fig. 1 illustrates the mortality and dialysis rates across income quartiles within each racial group. Panel A displays survey-weighted in-hospital mortality and panel B shows corresponding rates of dialysis use among adults hospitalized with AKI. Although modest socioeconomic gradients are observed within most racial groups, substantial racial differences persist across all income levels, indicating that income alone does not account for the observed disparities in AKI outcomes. Income quartiles correspond to ZIP code–level median household income.

### Adjusted AKI model

Adjusted odds of developing AKI are presented in Table 3. Compared with White patients, Black patients had significantly higher adjusted odds of AKI (OR 1.34, 95% CI 1.32–1.36), as did Native American (OR 1.08, 95% CI 1.03–1.13) and Other race patients (OR 1.07, 95% CI 1.05– 1.10). Asian/Pacific Islander patients had lower adjusted odds (OR 0.94, 95% CI 0.91–0.96), while Hispanic patients had similar odds (OR 0.98, 95% CI 0.96–1.00). CKD, sepsis, liver disease, heart failure, diabetes, and increasing age were the strongest predictors in the model.

**Table 3.** Adjusted Odds of Acute Kidney Injury Among Adult Hospitalizations.

| Predictor | OR | CI |
| --- | --- | --- |
| Asian/Pacific Islander | 0.937 | 0.913–0.962 |
| Black | 1.342 | 1.323–1.362 |
| Hispanic | 0.978 | 0.959–0.997 |
| Native American | 1.078 | 1.031–1.126 |
| Other | 1.070 | 1.046–1.096 |
| Primary payer | 1.017 | 1.013–1.021 |
| Age (per year) | 1.022 | 1.022–1.023 |
| Female | 0.678 | 0.673–0.684 |
| Chronic kidney disease | 3.849 | 3.788–3.911 |
| Heart failure | 1.454 | 1.44–1.467 |
| Diabetes | 1.342 | 1.329–1.355 |
| Hypertension | 1.091 | 1.081–1.101 |
| Obesity | 1.179 | 1.166–1.191 |
| COPD | 0.942 | 0.934–0.95 |
| Liver disease | 2.636 | 2.577–2.695 |
| Sepsis | 3.849 | 3.795–3.903 |

Table 3 shows that, after adjustment for demographics, comorbidities, and payer, Black, Native American, and Other race patients have higher odds of AKI, whereas Asian/Pacific Islander and (slightly) Hispanic patients have similar or lower risk; CKD, sepsis, liver disease, heart failure, diabetes, and older age are the strongest clinical predictors of AKI. Odds ratios represent the independent association of each predictor with the development of AKI during hospitalization. White race served as reference categories. Confidence intervals reflect survey-weighted standard errors.

### Adjusted mortality model

Adjusted odds of in-hospital mortality among AKI patients are shown in Table 4. Among AKI hospitalizations, Asian/Pacific Islander patients had higher adjusted odds of in-hospital mortality (OR 1.16, 95% CI 1.10–1.22) as did Native American (OR 1.19, 95% CI 1.07–1.31) and Other race patients (OR 1.17, 95% CI 1.11–1.23). In contrast, Black (OR 0.94, 95% CI 0.92–0.97) and Hispanic (OR 0.96, 95% CI 0.93–1.00) patients had slightly lower adjusted mortality compared with White patients. Sepsis, liver disease, heart failure, COPD, and older age were associated with higher adjusted mortality.

**Table 4.** Adjusted Odds of In-Hospital Mortality Among AKI Patients.

| Predictor | OR | CI |
| --- | --- | --- |
| Asian/Pacific Islander | 1.161 | 1.105–1.219 |
| Black | 0.942 | 0.915–0.97 |
| Hispanic | 0.961 | 0.927–0.996 |
| Native American | 1.186 | 1.07–1.314 |
| Other | 1.166 | 1.105–1.23 |
| Primary payer | 1.082 | 1.07–1.095 |
| Age (per year) | 1.018 | 1.017–1.019 |
| Female | 0.955 | 0.94–0.969 |
| Chronic kidney disease | 0.671 | 0.659–0.683 |
| Heart failure | 1.503 | 1.478–1.528 |
| Diabetes | 0.774 | 0.756–0.792 |
| Hypertension | 0.846 | 0.825–0.867 |
| Obesity | 0.731 | 0.714–0.748 |
| COPD | 1.058 | 1.037–1.079 |
| Liver disease | 3.005 | 2.905–3.108 |
| Sepsis | 4.652 | 4.561–4.745 |

Table 4 summarizes adjusted mortality risk among AKI patients, showing higher odds of in-hospital death for Asian/Pacific Islander, Native American, while Black and Hispanic patients have slightly lower adjusted mortality; sepsis, liver disease, heart failure, and older age are the dominant predictors of death. White race served as reference categories. Odds ratios indicate the independent association of each predictor with in-hospital death among AKI patients. Confidence intervals incorporate NIS sampling weights, strata, and hospital clustering.

### Adjusted dialysis model

Adjusted odds of dialysis among AKI patients appear in Table 5. All non-White racial and ethnic groups had higher adjusted odds of receiving dialysis compared with White patients, including Asian/Pacific Islander (OR 1.49, 95% CI 1.40–1.58), Hispanic (OR 1.34, 95% CI 1.28–1.41), Native American (OR 1.36, 95% CI 1.20–1.53), Other race (OR 1.30, 95% CI 1.20–1.40), and Black patients (OR 1.18, 95% CI 1.14–1.22). CKD, sepsis, liver disease, and heart failure were strongly associated with dialysis use. The adjusted associations for AKI, mortality, and dialysis are summarized visually in Figure 2, which displays all odds ratios and 95% confidence intervals in forest plot format.

**Table 5.** Adjusted Odds of Dialysis Among AKI Patients.

| Predictor | OR | CI |
| --- | --- | --- |
| Asian/Pacific Islander | 1.487 | 1.397–1.583 |
| Black | 1.180 | 1.14–1.221 |
| Hispanic | 1.343 | 1.28–1.408 |
| Native American | 1.355 | 1.204–1.525 |
| Other | 1.297 | 1.204–1.398 |
| Primary payer | 0.981 | 0.971–0.991 |
| Age (per year) | 0.971 | 0.971–0.972 |
| Female | 0.911 | 0.893–0.928 |
| Chronic kidney disease | 3.688 | 3.58–3.798 |
| Heart failure | 1.584 | 1.548–1.62 |
| Diabetes | 1.056 | 1.03–1.083 |
| Hypertension | 1.067 | 1.037–1.098 |
| Obesity | 1.023 | 0.996–1.05 |
| COPD | 0.872 | 0.848–0.897 |
| Liver disease | 2.253 | 2.134–2.379 |
| Sepsis | 2.477 | 2.414–2.542 |

**Figure 2.**
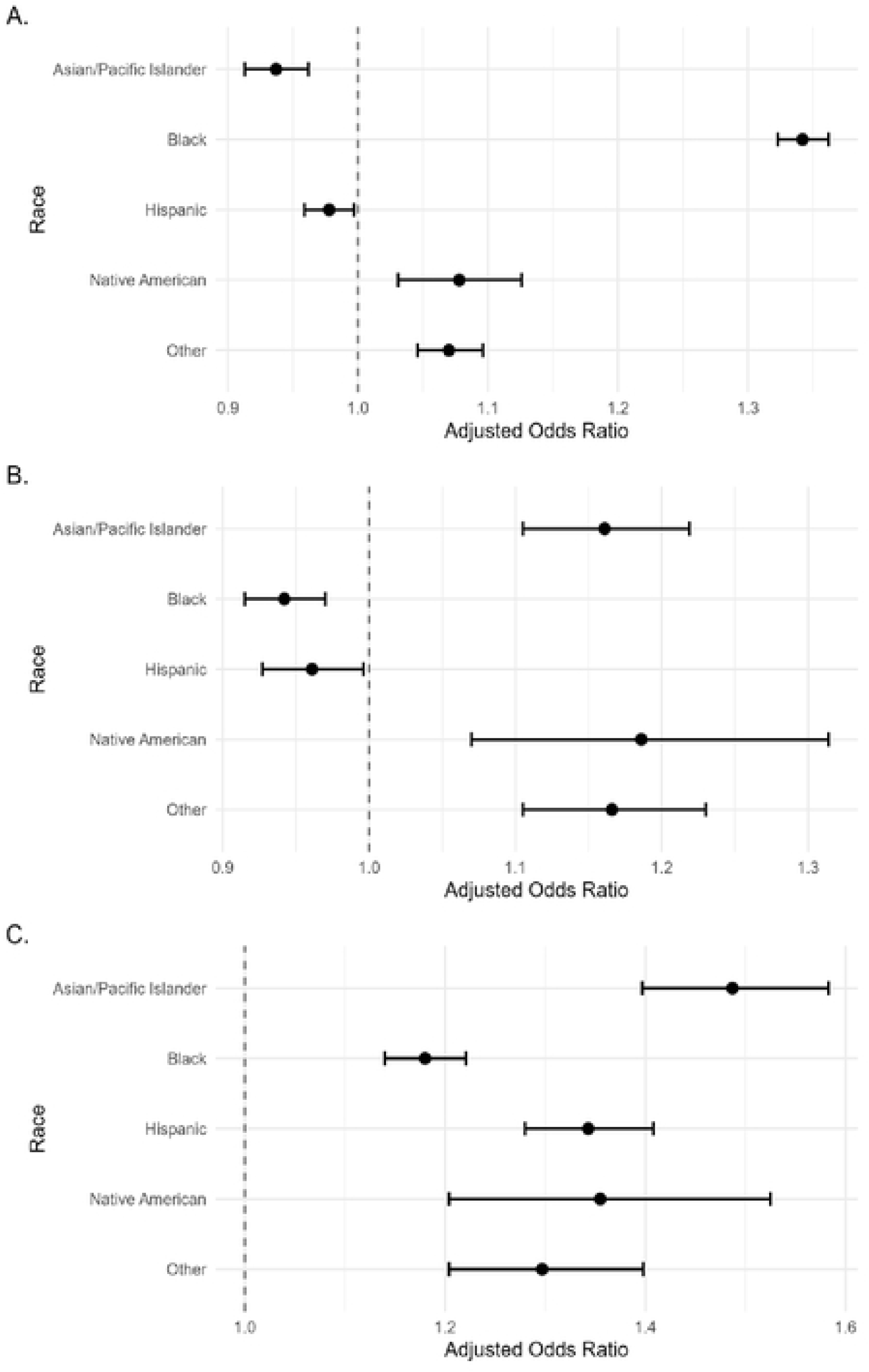
Adjusted Odds Ratios for AKI, In-Hospital Mortality, and Dialysis by Race.

Table 5 demonstrates that, among patients with AKI, all non-White racial groups have higher adjusted odds of requiring dialysis compared with White patients; CKD, sepsis, liver disease, and heart failure are the strongest clinical drivers of dialysis requirement, with younger and female patients somewhat less likely to receive dialysis. White race served as reference groups. The model reflects the independent association between each factor and the need for dialysis during the hospitalization. Confidence intervals were calculated using NIS survey design elements.

Fig. 2 illustrates adjusted racial disparities across all endpoints, showing differences in the odds of developing AKI, in-hospital mortality, and dialysis use when compared with White patients. Panel A presents adjusted odds ratios (ORs) for developing AKI among hospitalized adults; Panel B displays adjusted odds of in-hospital mortality among patients with AKI; and Panel C shows adjusted odds of receiving dialysis among AKI hospitalizations. White patients served as the reference group for all models. Error bars represent 95% confidence intervals. Models were adjusted for age, sex, primary payer, and major comorbidities including CKD, heart failure, diabetes, hypertension, obesity, COPD, liver disease, and sepsis.

### Age-Specific Predicted AKI Risk Differences

Modeled age-specific predicted AKI probabilities are shown in Figure 3. Across adulthood, Black patients consistently demonstrated higher predicted AKI probability than White patients, with absolute risk differences for Black patients reaching approximately 3–4 percentage points between ages 40 and 60 years, with smaller but consistently positive differences also observed for Native American and Other race patients. Asian/Pacific Islander patients showed lower predicted probabilities across most ages.

**Figure 3.**
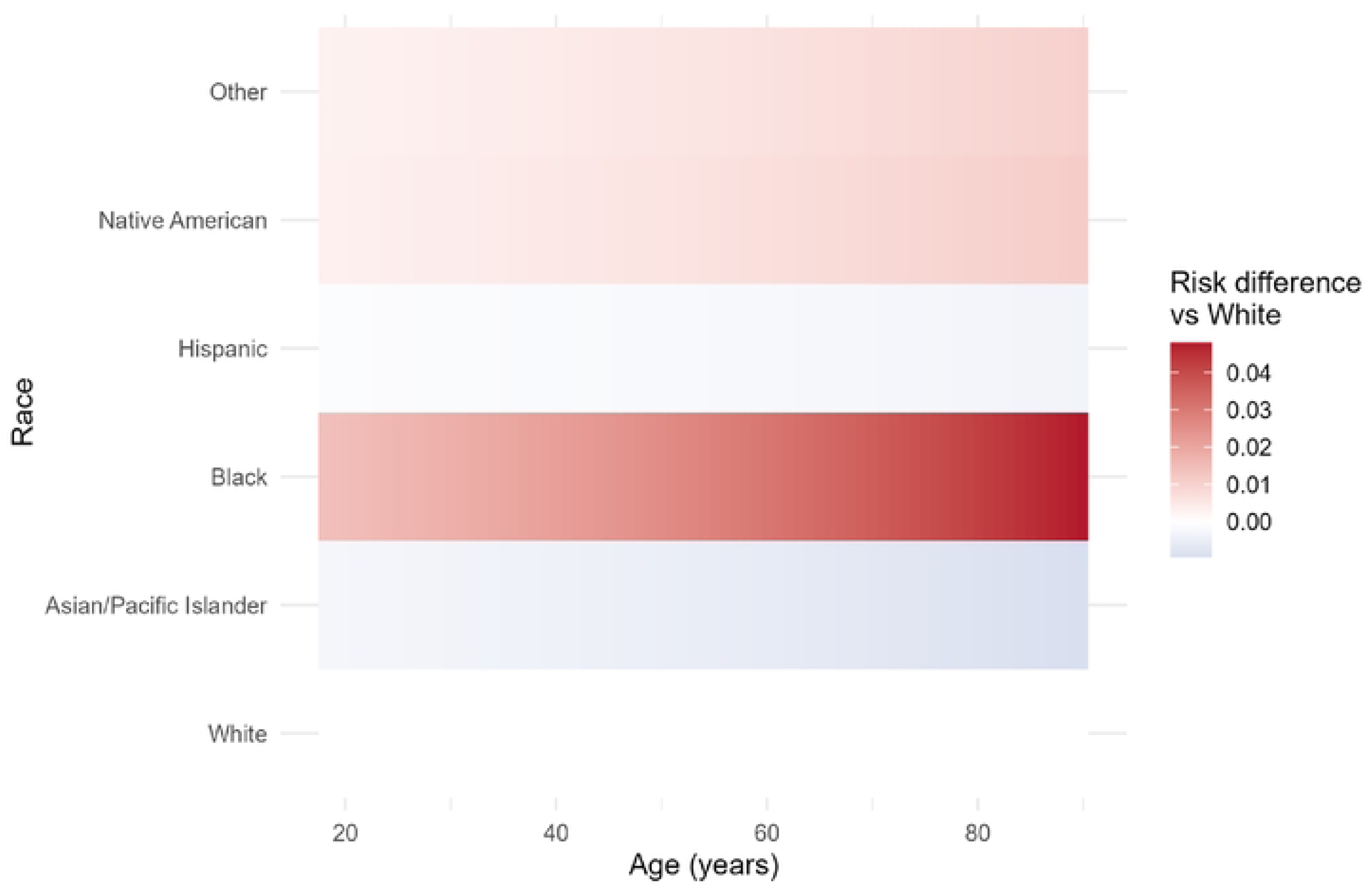
Risk Difference in Predicted AKI Probability Compared with Patients Across Age.

Fig. 3 illustrates the age-specific difference in predicted AKI probability between each racial group and White patients, demonstrating a consistently higher modeled risk for Black, Native American, and Other race patients and a lower predicted risk for Asian/Pacific Islander patients across the age spectrum. Risk differences represent absolute differences in predicted AKI probability relative to White patients, not adjusted odds ratios. Positive values (red) indicate higher predicted AKI probability relative to White patients of the same age, whereas negative values (gray) indicate lower predicted probability. Predicted probabilities were generated using the survey-weighted logistic regression model from Table 3. Covariates—including sex, primary payer, CKD, heart failure, diabetes, hypertension, obesity, COPD, liver disease, and sepsis—were held constant at reference values. Age was modeled as a continuous variable.

## Discussion

In this nationally representative analysis, we found substantial racial disparities in acute kidney injury and its downstream outcomes. Unadjusted differences in AKI prevalence, critical illness, and resource utilization were pronounced. These disparities remained after adjustment for demographics, payer status, and comorbidity profiles—consistent with prior AKI epidemiology and inequity research [6,8]. Figure 2 demonstrates that racial differences persisted despite extensive adjustment, and the age-specific predicted-probability curves in Figure 3 show that these inequities span the entire adult lifespan. Collectively, these findings highlight durable system-level inequities with important implications for kidney-health equity and inpatient care.

These inequities reflect not only clinical and comorbidity differences but also the long-standing influence of structural racism—manifested through concentrated neighborhood disadvantage, differential access to preventive and specialty kidney care, chronic underinvestment in minority-serving hospitals, and environmental and occupational exposures. Black, Native American, and Other race patients exhibited significantly higher adjusted odds of AKI, consistent with multiple national analyses documenting disproportionate AKI vulnerability in these groups [4,15–16]. While our dataset cannot directly measure structural contributors, prior literature suggests that upstream social and environmental determinants—including differential exposure to nephrotoxins, variability in preventive care, neighborhood disadvantage, and accumulated chronic disease burden—may help explain these patterns [1,8,20]. Asian/Pacific Islander patients demonstrated lower adjusted odds of AKI, which may relate to differences in risk profiles or other contextual factors described in population-based kidney studies [11]. The age-stratified risk curves reinforce that disparities are stable across the lifespan, suggesting cumulative exposure to inequities rather than age-specific drivers.

Racial patterns also extended to outcomes among patients who developed AKI. Prior research has described complex and sometimes paradoxical mortality patterns across racial and ethnic groups, potentially shaped by age distribution, survivorship, and hospital-level variation [9–10]. Our findings reflected this nuance. Black and Hispanic patients had slightly lower adjusted odds of in-hospital mortality, whereas Asian/Pacific Islander and Native American patients had higher odds (Figure 2). Similar paradoxical survival advantages among Black and Hispanic patients have been reported in AKI, sepsis, ESKD, and critical illness cohorts, and several proposed explanations exist.

Prior studies suggest that differences in pre-hospital health trajectories and selective survival into older adulthood may partly contribute to these patterns [9–10]. Additionally, some analyses of critically ill populations have proposed a “survivor bias,” wherein minority patients with severe chronic disease may be more likely to survive long enough to reach hospitalization, thereby appearing to have lower inpatient mortality despite higher long-term risk [6]. While our data cannot distinguish between these mechanisms, the observed pattern aligns with the broader literature and should be interpreted within the known limitations of administrative datasets.

Although income was associated with markers of illness severity and resource use, it did not explain the racial disparities observed. This is consistent with prior work showing that socioeconomic status captures only one dimension of structural inequity [17]. Factors such as segregation, healthcare accessibility, and hospital quality pathways reflect structural racism not fully measured by income alone.

Racial differences in resource utilization provide an additional dimension of inequity. Prior research has linked hospital variation and regional resource intensity to race-related differences in length of stay and cost [14]. Although length of stay differences in our study were modest, we found meaningful variation in hospital charges across racial and ethnic groups, with several minority groups incurring higher costs. Discharge disposition patterns also differed. Minority patients were less likely to transition to skilled nursing facilities and more likely to leave against medical advice, consistent with studies attributing these trends to barriers in post-acute care access, unmet social needs, and challenges in continuity of care [5]. Collectively, these utilization patterns may contribute to downstream kidney outcomes and broader health inequities.

To meaningfully address these inequities, policy efforts must target the structural and institutional drivers of AKI risk. Strategies include expanding equitable access to early nephrology consultations, particularly in minority-serving hospitals where specialty availability is often limited; implementing hospital-level quality audits to identify and remediate disparities in AKI recognition, fluid management, and dialysis initiation; and improving outpatient CKD surveillance in minoritized communities through community health worker programs, mobile clinics, and enhanced primary care-nephrology coordination. Investments in upstream social and community infrastructure, including improved access to clean water, reduction of environmental nephrotoxins, and transportation support for medical visits, are also essential. These interventions require multisector collaboration and sustained resource allocation, reflecting the reality that racial disparities in AKI stem from long-standing structural racism embedded across healthcare, housing, and economic systems.

This study has several strengths. The National Inpatient Sample provides national generalizability across diverse populations and hospital settings. Use of survey weights and multivariable adjustment strengthens internal validity. The inclusion of multiple endpoints—AKI incidence, mortality, dialysis requirement, resource utilization, and discharge disposition—allows a multidimensional characterization of inequity.

Several limitations merit discussion. Administrative coding may misclassify AKI, particularly milder presentations, compared with laboratory-based KDIGO definitions. The absence of present-on-admission indicators limits temporal inference regarding comorbidities and complications. Important clinical variables—including nephrotoxin exposure, fluid status, baseline kidney function, and urine output—are unavailable in administrative datasets. Race is administratively recorded and may not capture sociocultural or structural exposures. ZIP code–level income is an imperfect proxy for individual socioeconomic position. These limitations likely bias estimates toward the null, suggesting that true disparities may be even greater than observed.

In summary, this nationwide analysis demonstrates persistent racial disparities in AKI risk, mortality, dialysis utilization, and resource use. These inequities were not fully explained by comorbidities or socioeconomic factors and were evident across the adult lifespan. Efforts to reduce disparities in AKI care will require targeted prevention strategies, equitable access to nephrology services, and policy-level interventions that address upstream social and structural drivers of kidney disease. A coordinated clinical and public health response will be essential to meaningfully improve kidney-health equity at a national scale.

## Data Availability

The data that support the findings of this study are available from the Agency for Healthcare Research and Quality, Department of Health and Human Services of the United States. However, restrictions apply to the availability of these data, which were used under license for the current study, and so are not publicly available. Data are, however, available from the corresponding author upon reasonable request and with permission of the Agency for Healthcare Research and Quality.

http://hcup-us.ahrq.gov/db/hcupdatapartners.jsp

## Acknowledgements

We wanted to acknowledge all the HCUP Data Partners that contribute to HCUP. A link to the HCUP-US web page that contains the list of State organizations is here. (<u>hcup-us.ahrq.gov/db/hcupdatapartners.jsp</u>).

We thank Patryk Klimek, clinical project manager at BayCare Health System, for his helpful contributions to manuscript revisions.

## Notes

### Competing Interest Statement

NO authors have competing interests

### Funding Statement

The authors declare that they received no funding.

### Author Declarations

This project was reviewed by the BayCare Institutional Review Board (IRB) and determined not to constitute human subjects research as defined by DHHS and FDA regulations (Inquiry determination dated October 22, 2025). IRB approval and oversight were therefore not required. The analysis used fully de-identified data from the Healthcare Cost and Utilization Project (HCUP) National Inpatient Sample, and no identifiable private information or intervention involving human participants occurred. All study procedures were conducted in accordance with the ethical principles of the Declaration of Helsinki. Because no human subjects were involved, informed consent was not required.

## References

1. Berry J, Perez A, Di M, Hu C, Pastan SO, Patzer RE, Harding JL. The Association Between Residential Segregation and Access to Kidney Transplantation–Evidence from a Multi-State Cohort Study. Clinical Journal of the American Society of Nephrology. 2024 Jan 24:10–2215.

2. Boulware LE, Mohottige D. The seen and the unseen: race and social inequities affecting kidney care. Clinical Journal of the American Society of Nephrology. 2021 May 1;16(5):815–7.

3. Chertow GM, Burdick E, Honour M, Bonventre JV, Bates DW. Acute kidney injury, mortality, length of stay, and costs in hospitalized patients. Journal of the American Society of Nephrology. 2005 Nov 1;16(11):3365–70.

4. Evans K, Coresh J, Bash LD, Gary-Webb T, Köttgen A, Carson K, Boulware LE. Race differences in access to health care and disparities in incident chronic kidney disease in the US. Nephrology Dialysis Transplantation. 2011 Mar 1;26(3):899–908.

5. Harrison TG, Scory TD, Hemmelgarn BR, Brindle ME, Daodu OO, Graham MM, James MT, Lam NN, Roshanov P, Sauro KM, Ronksley PE. Differences in Postoperative Disposition by Kidney Disease Severity: A Population-Based Cohort Study. American Journal of Kidney Diseases. 2025 May 1;85(5):589–602.

6. Hassan MO, Owoyemi I, Abdel-Rahman EM, Ma JZ, Balogun RA. Association of race with in-hospital and post-hospitalization mortality in patients with acute kidney injury. Nephron. 2021 May 5;145(3):214–24.

7. HCUP National Inpatient Sample (NIS). Healthcare Cost and Utilization Project (HCUP). 2022. Agency for Healthcare Research and Quality, Rockville, MD. hcup-us.ahrq.gov/nisoverview.jsp

8. Hounkpatin HO, Fraser SD, Honney R, Dreyer G, Brettle A, Roderick PJ. Ethnic minority disparities in progression and mortality of pre-dialysis chronic kidney disease: a systematic scoping review. BMC nephrology. 2020 Jun 9;21(1):217.

9. Kalantar-Zadeh K, Kovesdy CP, Derose SF, Horwich TB, Fonarow GC. Racial and survival paradoxes in chronic kidney disease. Nature clinical practice Nephrology. 2007 Sep;3(9):493–506.

10. Kalantar-Zadeh K, Kovesdy CP, Norris KC. Racial survival paradox of dialysis patients: robust and resilient. American journal of kidney diseases: the official journal of the National Kidney Foundation. 2012 Apr 10;60(2):182.

11. Kataoka-Yahiro M, Davis J, Gandhi K, Rhee CM, Page V. Asian Americans & chronic kidney disease in a nationally representative cohort. BMC nephrology. 2019 Jan 9;20(1):10.

12. Lafrance JP, Miller DR. Acute kidney injury associates with increased long-term mortality. Journal of the American Society of Nephrology. 2010 Feb 1;21(2):345–52.

13. Lapedis CJ, Mariani LH, Jang BJ, Hodgin J, Hicken MT. Understanding the link between neighborhoods and kidney disease. Kidney360. 2020 Aug 1;1(8):845–54.

14. Mathioudakis NN, Giles M, Yeh HC, Haywood Jr C, Greer RC, Golden SH. Racial differences in acute kidney injury of hospitalized adults with diabetes. Journal of Diabetes and its Complications. 2016 Aug 1;30(6):1129–36.

15. Mehrotra R, Kermah D, Fried L, Adler S, Norris K. Racial differences in mortality among those with CKD. Journal of the American Society of Nephrology. 2008 Jul 1;19(7):1403–10.

16. Narva AS. Reducing the burden of chronic kidney disease among American Indians. Advances in chronic kidney disease. 2008 Apr 1;15(2):168–73.

17. Nicholas SB, Kalantar-Zadeh K, Norris KC. Socioeconomic disparities in chronic kidney disease. Advances in chronic kidney disease. 2015 Jan 1;22(1):6–15.

18. Norton JM, Moxey-Mims MM, Eggers PW, Narva AS, Star RA, Kimmel PL, Rodgers GP. Social determinants of racial disparities in CKD. Journal of the American Society of Nephrology. 2016 Sep 1;27(9):2576–95.

19. Patzer RE, McClellan WM. Influence of race, ethnicity and socioeconomic status on kidney disease. Nature Reviews Nephrology. 2012 Sep;8(9):533–41.

20. Soderland P, Lovekar S, Weiner DE, Brooks DR, Kaufman JS. Chronic kidney disease associated with environmental toxins and exposures. Advances in chronic kidney disease. 2010 May 1;17(3):254–64.

21. Vart P, Gansevoort RT, Joosten MM, Bültmann U, Reijneveld SA. Socioeconomic disparities in chronic kidney disease: a systematic review and meta-analysis. American journal of preventive medicine. 2015 May 1;48(5):580–92.

22. Vart P, van Zon SK, Gansevoort RT, Bültmann U, Reijneveld SA. SES, chronic kidney disease, and race in the US: a systematic review and meta-analysis. American journal of preventive medicine. 2017 Nov 1;53(5):730–9.

23. Venkatachalam MA, Griffin KA, Lan R, Geng H, Saikumar P, Bidani AK. Acute kidney injury: a springboard for progression in chronic kidney disease. American Journal of Physiology-Renal Physiology. 2010 May;298(5):F1078–94.

24. Zarkowsky DS, Arhuidese IJ, Hicks CW, Canner JK, Qazi U, Obeid T, Schneider E, Abularrage CJ, Freischlag JA, Malas MB. Racial/ethnic disparities associated with initial hemodialysis access. JAMA surgery. 2015 Jun 1;150(6):529–538.

